# Estimating Population Counts for Dissemination Areas and Census Tracts in Canada from 2011 to 2021

**DOI:** 10.1101/2025.02.28.25322945

**Authors:** Anousheh Marouzi, Charles Plante

## Abstract

Accurate small-area population estimates are essential for health research and social policy development. While Statistics Canada provides census-year population counts for all geographic units, it does not produce intercensal estimates for Dissemination Areas (DA) and Census Tracts (CT). This study addresses this gap by estimating DA- and CT-level population counts for 2011 to 2021 using census data and interpolation techniques. Population counts were derived from Statistics Canada’s Geographic Attribute Files for 2011, 2016, and 2021. We applied linear interpolation to estimate intercensal population counts (2012–2015 and 2017–2020). We used an areal-weighted interpolation technique to account for boundary shifts due to census geography changes, utilizing Statistics Canada’s Correspondence Files. The final datasets provide consistent population estimates across census cycles, enabling longitudinal and neighbourhood-level analyses. The methodology and accompanying R script, available as supplementary materials, can be adapted for other intercensal periods and other demographic information, promoting transparency and reproducibility in demographic research. This study facilitates data-driven decision-making in public health and policy development by providing a reliable and scalable methodology for estimating intercensal population counts.

**About the Research Department:** The Saskatchewan Health Authority Research Department leads collaborative research to enhance Saskatchewan’s health and healthcare. We provide diverse research services to SHA staff, clinicians, and team members, including surveys, study design, database development, statistical analysis, and assistance with research funding. We also spearhead our own research programs to strengthen research and analytic capability and learning within Saskatchewan’s health system.

**Disclaimer:** This working paper is for discussion and comment purposes. It has not been peer-reviewed nor been subject to review by Research Department staff or executives. Any opinions expressed in this paper are those of the author(s) and not those of the Saskatchewan Health Authority.

**Suggested Citation:** Marouzi Anousheh, Plante Charles. 2025. “Estimating Population Counts for Dissemination Area and Census Tracts in Canada from 2011 to 2021.” MedRxiv.

**Author Contributions:** AM conducted the data analysis and prepared the first draft of the article. AM and CP designed the study and directed its implementation, including quality assurance and control. CP supervised the data analysis. CP reviewed, edited, and finalized the text. CP provided the overall guidance and funding for the research project. All authors approved the final version of the manuscript.

**Funding Statement:** This research was funded by the Saskatchewan Health Research Foundation (SHRF).

**Ethics Declaration:** This study exclusively utilizes publicly available, de-identified population data obtained from Statistics Canada. No human participants, personal identifiers, or confidential information were involved in this research, and therefore, ethical approval was not required.

**Conflict of Interest:** The authors declare that they have no conflict of interest.

**Data Availability:** All data used in this study is for public use and can be accessed through the Statistics Canada website.

**Code Availability:** Codes are available as a supplementary file to this working paper.

## Background

Population counts are a cornerstone of health research and social policy development, providing the foundation for calculating key indicators, tracking demographic trends, and assessing disparities. Statistics Canada collects demographic data on the Canadian population through a national Census of Population conducted every five years. Census-year population counts are publicly available on Statistics Canada’s website for all census geographies, but, since the census is conducted only every five years, the counts for intercensal years are not directly available. To address this data gap, Statistics Canada has developed a methodology for estimating intercensal population counts at the Census Subdivision (CSD), Census Division (CD), Census Metropolitan Area (CMA), Census Agglomeration (CA), and provincial/territorial levels^1^ and report these estimates publicly.^2,3^ However, intercensal population estimates are not provided for the smaller geographic units of Census Tracts (CTs) and Dissemination Areas (DAs).

This gap poses challenges for local-level studies that use CTs and DAs as proxies or component pieces of small geographic areas, such as for neighbourhoods or rural towns and communities. CTs and DAs are widely used in health and social epidemiology studies due to their relatively small size and socioeconomic homogeneity. These geographic units are instrumental in analyzing health outcomes and inequalities at the neighbourhood level.^4–6^ Additionally, several Canadian deprivation indices are constructed at these levels, reinforcing the demand for accurate population counts at that level.^7–10^ Since intercensal data for DAs and CTs are not readily accessible, researchers commonly devise and employ their own interpolation techniques to fill the data gap. This can be challenging, especially for new researchers, as census boundaries are revised every five years to reflect demographic changes and Statistics Canada does not provide census and geographic attribute files needed for interpolations in a single location.

The primary objective of this study is to provide researchers with open-source DA- and CT-level population counts for all years between 2011 and 2021 by collating disparate counts provided by Statistics Canada and interpolating counts for intercensal years. Furthermore, this study details the methodology and provides an R script (Appendix A) to facilitate reproducibility, allowing researchers to adapt the approach for different intercensal periods or small-area demographic analyses.

## Methods

### Data Sources

Population counts were derived from Statistics Canada’s Geographic Attribute Files for the 2011, 2016, and 2021 censuses.^11^ These files provide official population counts at the Dissemination Block (DB) level, the smallest geographic unit for which population and dwelling counts are disseminated.^12,13^ Correspondence Files from 2016 and 2021 were incorporated to address changes in geographic boundaries. These files detail geographic overlap between dissemination areas among adjacent censuses.^14,15^ Geographic boundaries in Canada are updated by Statistics Canada with every census; therefore, census geographies are not the same between censuses.

### Geographic Units

This study estimates population counts for DAs and CTs, geographic units for which Statistics Canada does not provide intercensal population estimates. DAs represent the smallest census units for which demographic and socioeconomic data are publicly provided by Statistics Canada in its Census Profiles and typically contain between 400 and 700 individuals.^16,17^ Census Tracts are larger geographic units, generally encompassing populations of 2,500 to 7,500 residents, and are delineated only within larger CMAs and CAs.^18,19^

CTs are particularly useful for analyzing socioeconomic conditions at the neighbourhood level. Their size closely corresponds to city neighbourhoods, and Statistics Canada delineates them to ensure homogeneity in socioeconomic characteristics such as income levels and social living conditions.^18,19^ Since Statistics Canada does not provide direct intercensal population estimates for DAs or CTs, we developed an estimation approach that integrates linear interpolation for temporal continuity and areal-weighted methods to account for geographic boundary changes.

### Population count estimation, 2011 to 2021

#### Census year population counts

For the 2011, 2016, and 2021 census years, DA- and CT-level population counts were calculated by aggregating DB population data from Statistics Canada’s Geographic Attribute Files.^11^ Statistics Canada applies random rounding to base-5 at the DB level to protect confidentiality.

An internal consistency procedure ensures that aggregated counts at lower geographic levels, such as DBs, sum accurately to higher geographic levels, including DAs and CTs.^13^ This verification process helps maintain consistency across geographic scales, minimizing distortions introduced by random rounding. The resulting DA and CT population counts provide a stable foundation for subsequent interpolation methods used to estimate intercensal populations.

#### Intercensal population estimates

To estimate populations for intercensal years (2012–2015, 2017–2020), we applied linear interpolation, which assumes that population growth follows a linear function of time between two census years and has previously been incorporated in the estimation of intercensal demographic data.^1,20^ For example, using this approach to interpolate between the 2011 and 2016 censuses, we calculate the population counts for intercensal periods *p*_*t*_ where *t* ∈ {2012, 2013, 2014, 2015} as:

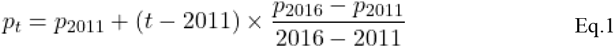

where *p*_*2011*_ and *p*_*2016*_ are the known population counts from census years 2011 and 2016, respectively. Since DA boundaries are nested within CT boundaries, we derived CT-level population estimates by aggregating interpolated DA counts. For consistency, all interpolated values were rounded to the nearest integer.

#### Adjusting for boundary changes between censuses

The linear interpolation approach in Eq.1 assumes no change in boundaries between censuses; however, census boundaries are updated with every census to reflect demographic changes, resulting in changes such as the splitting, merging, or reconfiguring of DAs. Before employing this approach, we must geographically harmonize population counts between censuses. Taking 2011 and 2016 as our examples again, we can do so by calculating population counts for 2016 based on 2011 geographies. We do so using areal-weighted interpolation.^21,22^ This is a convenient approach since areal-overlap percentages are provided by Statistics Canada to support this kind of interpolation in their Correspondence Files.^15,23^

To estimate DA population counts in 2016 based on the boundaries of the previous census, 2011, *p*_*2016*↦*2011*_, we used the following equation:

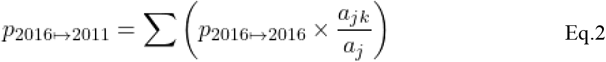

where *p*_*2016*↦*2016*_ is the population of DA *j* in 2016 based on its 2016 boundaries, *a*_*jk*_ is the land area of DA *j* in 2016 that overlaps with the land area of a target DA *k* in 2011, and *a*_*j*_ is the total land area of DA *j* in 2016.

We estimated population counts for 2012 to 2015 based on 2011 boundaries, that is using *p*_*2011*↦*2011*_ and *p*_*2016*↦*2011*_, and for 2017 to 2020 based on 2016 boundaries (see Figure 1), that is using *p*_*2016*↦*2016*_ and *p*_*2021*↦*2016*_. We only interpolated forwards and only for five-year intervals because census geographies change over time. While it can be tempting to hold geographies constant over time, this is nonsensical over longer time spans. Consider, for example, the metro area of Toronto, which has expanded by hundreds of DAs between 2006 and 2021, from 7,012 to 7,716. Neither are the boundaries of Toronto in 2006 meaningful in 2021, nor are the boundaries of Toronto in 2021 meaningful in 2006.

**Figure 1.**
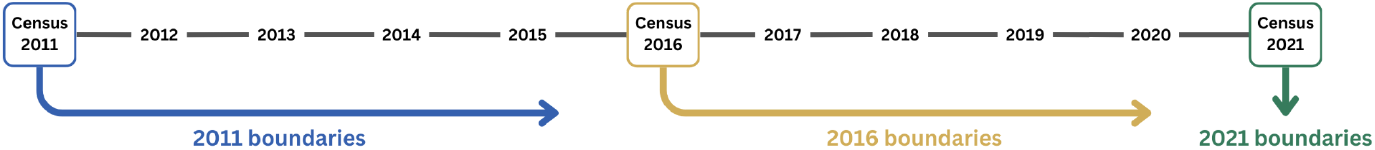
Boundaries used to report population counts for census and intercensal years.

Although updating boundaries at every census may be somewhat arbitrary in the short term, it ensures the alignment of areal units and meaningful geographic concepts like “Toronto” over the long term. It is also consistent with Statistics Canada’s practices. Moreover, Statistics Canada does not provide Correspondence Files for non-adjacent censuses.

### Output and recommendations for use

The final dataset consists of five comma-separated value (.CSV) files containing DA- and CT-level population estimates from 2011 to 2021. Table 1 provides an overview of the structure of these files. The geographic units used for reporting population counts are illustrated in Figure 1. Researchers can integrate these population count tables into their studies by merging them using the DA unique identifier variable (da_id_‘year’) included in the CSV files. The R script used to generate these estimates is provided in Appendix A, allowing researchers to replicate and extend the methodology for additional years or similar applications in demographic research.

**Table 1.** Data products for DA and CT population counts in Canada from 2011 to 2021.

| **Dataset name** | **Years** | **Data level** | **Method of calculation\*** |
| --- | --- | --- | --- |
| pop_counts_da11_2011_RELEASE.csv | 2011 | DA 2011 | Summation of DB population counts |
| pop_counts_da11_2012to2015_RELEASE.csv | 2012 to 2015 | DA 2011 | Interpolation of DB population counts |
| pop_counts_da16_2016_RELEASE.csv | 2016 | DA 2016 | Summation of DB population counts |
| pop_counts_da16_2017to2020_RELEASE.csv | 2017 to 2020 | DA 2016 | Interpolation of DB population counts |
| pop_counts_da21_2021_RELEASE.csv | 2021 | DA 2021 | Summation of DB population counts |

Note: DA = Dissemination Area, CSD = Census Subdivision, CT = Census Tract, DB = Dissemination Block. \*Population counts produced by summation of DB counts (as reported in Statistics Canada Attribute Files) are true values, and counts produced by the interpolation technique are estimates.

The areal-weighted interpolation method we used solely redistributes population counts based on the proportional overlap between census geographies and, therefore, assumes a uniform population distribution within each geographic unit. Researchers may wish to enhance the accuracy of their estimates by leveraging more advanced GIS-based areal interpolation techniques that have been developed in Canada and the United States to address this limitation.^18,24,25^ For example, the Canadian Longitudinal Census Tract Database (CLTD) developed by Allen and Taylor^18^ employs a combination of map-matching techniques, including dasymetric overlays and population-weighted areal interpolation, to produce apportionment tables bridging CT-level data for the quinquennial census from 1971 to 2020.^1^

It is important to note that the intercensal population counts produced in this study are estimates and not exact values. Previous research has demonstrated that linear interpolation provides estimates with relatively small errors for population counts, particularly in more populous areas.^20^ However, researchers should carefully evaluate whether the level of error associated with this method, as discussed by Weden et al.,^20^ is acceptable for their specific research questions.

## Conclusion

This paper presents an approach to estimating intercensal population counts for small geographic units in Canada, specifically at the DA and CT levels. It provides a helpful tool for longitudinal and small-area demographic analysis by utilizing publicly available census data and interpolation methods. While the methodology produces reasonable estimates, it has limitations, particularly in areas with rapid demographic changes. Nevertheless, in the absence of official intercensal estimates for small geographic units, this dataset is likely to serve as an essential resource for health research and policy development.

As mentioned above, future research could refine the interpolation techniques in our study by incorporating the population-weighted apportionment methods and dasymetric overlays to enhance the accuracy recommended by Allen and Taylor.^18^ Their methodology, however, does necessitate a basic facility with GIS software. Improving estimation methodologies will support more precise and data-driven decision-making in health and social sciences.

Statistics Canada’s Population and Family Estimation Methods offer more robust estimates for larger geographic areas by incorporating factors such as birth, death, immigration, and emigration.^1^ We advise that researchers working on larger areas not aggregate our estimates but rather use these official estimates. We advise only using the methodology presented here at finer spatial scales where no alternative exists.

## Supporting information

Population Counts 2011 to 2021

## Data Availability

All data used in this study is for public use and can be accessed through the Statistics Canada website.

## Acknowledgments

We are thankful to Dr. Jef Allen and Dr. Zack Taylor for the valuable information on the Canadian Longitudinal Census Tract Database (CLTD). We also thank Dr. Suvadra Datta Gupta for reviewing the final version of the current paper and for her helpful comments. All views expressed in this work are our own.

## Footnotes

1 These tables are publicly available at https://github.com/jamaps/CLTD/tree/master.

