## Appendix A for "Estimating Population Counts for Dissemination Areas and Census Tracts in Canada from 2011 to 2021": 2024SHRF_population counts_2011to2021_202501_02.r

```
################################################################################
#
# Copyright © 2025 Authors
# 
# This code is distributed under the Creative Commons Attribution 4.0 
# International License (CC BY 4.0).
# 
# By using this code, you agree to:
#
#   1. Cite the associated publication: 
#
#      Marouzi Anousheh, Plante Charles. "Estimating Population Counts for 
#      Dissemination Areas and Census Tracts in Canada from 2011 to 2021,"
#      medRxiv, 2025. 
#
#   2. If you share this code or derivatives, include this copyright and terms 
#      of use statement. 
#
#   3. This code is provided "as-is," without warranty of any kind. 
# 
#
################################################################################

# ESTIMATING POPULATION COUNTS FOR DA AND CT IN CANADA FROM 2011 TO 2021    ####
# ============================================================================ #
# Summary: This R script calculates population counts for years 2011, 2016,
# and 2021 at various geographic levels, using the attribute files publicly 
# provided by Statistics Canada. Additionally, it estimates population counts 
# for intercensal years by linear interpolation.
#
# Programmer: Anousheh Marouzi
#
# Start date: January 10th, 2025
# 
# Last update: January 17th, 2025
# ============================================================================ #

# FILE NAMING CONVENTIONS                                                   ####
# ---------------------------------------------------------------------------- #
# The purpose of this R Script is to manipulate and link different datasets to 
# create a data product (i.e. CASDOHI) that can be used by other researchers. 
# Each section opens a dataset, manipulates it, and saves it by a meaningful 
# name. 
#
# In the current R Script, we name datasets according to a set of rules to 
# facilitate understanding of what they contain, how they relate to other 
# files, and the process a dataset goes through. 
#
# An overview of the structure of a dataset file's name is: 
#
#  'main information'_'geo unit'_`years covered'_`stage'
#
# Each section of this structure is explained below:
#
# 1. Main information: This part of the name indicates the type of 
#    information included in the file. 
#
# 2. Geo unit: This indicates the level of data. For example, da11 means 
#    that the dataset is provided at Dissemination Area 2011 level.
#
# 3. Years covered: This indicates the years that are covered in the dataset.
#
# 4. Stage: Files in this do-file are classified into three main groups: 
#    Raw, Master, and Use. These are indicative of three main steps that a 
#    dataset usually goes through in this do-file; A dataset matures from 
#    being a Raw file into  a Master file and finally becomes a Use file. 
#    However, these are not the  only three statuses of a dataset. Between 
#    the Raw and Master steps, we may save the dataset as a Reduced or 
#    Harmonized file. Following is a flow chart of how a dataset is processed 
#    in this do-file: 
#    
#         RAW --> (REDUCED) --> (HARM) --> MASTER --> USE --> RELEASE
#    
#    We add a suffix to the name of a dataset to show these stages. Below, we 
#    describe these 5 stages and provide the suffix in parentheses as they  
#    exist in the do-file:
#
#      a. Raw (_RAW): The first step in this do-file is to rename all raw  
#         datasets files according to our naming structure. This is done to    
#         have a consistent and meaningful name convention in our do-files 
#         across all our projects. We put the suffix "_RAW" at the end of the 
#         renamed raw files. These files are 100% similar to the original ones 
#         as provided in RDC.
#    
#      b. Reduced (_REDUCED): We add this suffix to datasets that we have 
#         dropped some of its variables or observations (reduced the number of 
#         variables or observations.) This step is usually done to speed up  
#         run times.
#     
#      c. Harmonized (_HARM): This suffix is added to datasets when its
#         variables are manipulated in a way to be harmonized in relation to
#         other datasets in the do-file. For example, variable sex is recorded
#         in different datasets to be consistent with how it is coded in the 
#         Census.
#     
#      d. Master (_MASTER): The Master stage of a dataset happens when general 
#         manipulations, such as reducing the number of variables and recoding
#         them, have been done, and the dataset is ready for more specialized
#         manipulation. A key feature of a Master file is that it usually 
#         contains the same number of rows as the raw file.
#    
#      e. Use (_USE): We add the suffix "USE" to a dataset when we have 
#         manipulated the Master dataset in a way that is ready for analysis.
#         This analysis can be descriptive, modeling, or any other type of 
#         analysis that produces the results of a research project. This step 
#         usually involves generating variables and dropping unnecessary 
#         observations.
#
#      f. Release (_RELEASE): The suffix "RELEASE" is added to the files that
#         are going to be released from RDC (in RDC context) or publish for 
#         public use (in outside-RDC context).
#
#
# ---------------------------------------------------------------------------- #

# LIST OF DATASETS IN THIS R SCRIPT                                         ####
# ---------------------------------------------------------------------------- #
# Inputs:
#   1. attribute_file_db11_2011_RAW.txt
#   2. attribute_file_db16_2016_RAW.csv
#   3. attribute_file_db21_2021_RAW.csv
#   4. correspondence_file_2016_RAW.csv
#   5. correspondence_file_2021_RAW.csv
#
# Outputs:
#   1. attribute_file_da_2011_MASTER.csv
#   2. attribute_file_da16_2016_MASTER.csv
#   3. attribute_file_da21_2021_MASTER.csv
#   4. correspondence_file_2016_HARM.csv
#   5. correspondence_file_2021_HARM.csv
#   6. pop_counts_da11_2016_MASTER.cs
#   7. pop_counts_da16_2021_MASTER.csv
#   8. pop_counts_da11_2011_RELEASE.csv
#   9. pop_counts_da11_2012to2015_RELEASE.csv
#   10. pop_counts_da16_2016_RELEASE.csv
#   11. pop_counts_da16_2017to2020_RELEASE.csv
#   12. pop_counts_da21_2021_RELEASE.csv
#
# ---------------------------------------------------------------------------- #

# TABLE OF CONTENTS                                                         ####
# ---------------------------------------------------------------------------- #
# 0. SETUP
# 1. CALCULATE 2011, 2016, AND 2021 POPULATION COUNTS
#   1.1. HARMONIZE 2011 ATTRIBUTE FILE AND CALCULATE POP COUNTS
#   1.2. HARMONIZE 2016 ATTRIBUTE FILE AND CALCULATE POP COUNTS
#   1.3. HARMONIZE 2021 ATTRIBUTE FILE AND CALCULATE POP COUNTS
# 2. HARMONIZE CORRESPONDENCE FILES - 2016 AND 2021
#   2.1. HARMONIZE 2016 CORRESPONDENCE FILE
#   2.2. HARMONIZE CORRESPONDENCE FILE, 2021
# 3. JOIN HARMONIZED ATTRIBUTE AND CORRESPONDENCE FILES - 2016 AND 2021
#   3.1 JOIN ATTRIBUTE AND CORRESPONDENCE - 2016
#   3.2 JOIN ATTRIBUTE AND CORRESPONDENCE - 2021
# 4. CALCULATE POPULATION COUNTS 
#   4.1. CALCULATE POP COUNTS FOR 2011 TO 2016 BASED ON 2011 GEO
#   4.2. CALCULATE POP COUNTS BETWEEN 2016 AND 2021 BASED ON 2016 GEO
#
# ---------------------------------------------------------------------------- #

# VERSION 1                                                                 ####
# ---------------------------------------------------------------------------- #
# This is the first version of the file.
# ---------------------------------------------------------------------------- #

# 0. SETUP                                                                  ####
# ---------------------------------------------------------------------------- #
# Clear environment
rm(list=ls(all=TRUE)) 

# Call packages
library(tidyverse)
library(stringr)
library(janitor)

# Set directory
setwd("/Users/Anousheh/Desktop/Research/CASDOHI")
# ---------------------------------------------------------------------------- #

# 1. CALCULATE 2011, 2016, AND 2021 POPULATION COUNTS                       ####
# ---------------------------------------------------------------------------- #
# Here, we harmonize the attribute files available on StatsCan website and
# calculate populations at different geographic levels for each census year.

  # 1.1. HARMONIZE 2011 ATTRIBUTE FILE AND CALCULATE POP COUNTS             ####
  # -------------------------------------------------------------------------- #

  # Define column positions in the raw attribute file according to the codebook
  # to import the txt file in the next step
  col_positions <- fwf_positions(
  
    # Define where variables start
    start = c(1, 11, 19, 27, 35, 48, 49, 57, 74, 91, 100, 111, 113, 168, 198, 
              228, 238, 248, 253, 338, 342, 427, 431, 471, 474, 481, 536, 539, 
              540, 543, 550, 605, 611, 696, 699, 704, 707, 807, 808, 818, 822, 
              829, 835, 839, 939, 940),
  
    # Define where variables end 
    end = c(10, 18, 26, 34, 47, 48, 56, 73, 90, 99, 110, 112, 167, 197, 227, 
            237, 247, 252, 337, 341, 426, 430, 470, 473, 480, 535, 538, 539, 
            542, 549, 604, 610, 695, 698, 703, 706, 806, 807, 817, 821, 828, 
            834, 838, 938, 939, 941),
  
    # Define variable names
    col_names = c("DBuid", "DBpop2011", "DBtdwell2011", "DBurdwell2011", 
                  "DBarea", "DB_ir2011", "DAuid", "DAlamx", "DAlamy", "DAlat", 
                  "DAlong", "PRuid", "PRname", "PRename", "PRfname", "PReabbr", 
                  "PRfabbr", "FEDuid", "FEDname", "ERuid", "ERname", "CDuid", 
                  "CDname", "CDtype", "CSDuid", "CSDname", "CSDtype", "SACtype", 
                  "SACcode", "CCSuid", "CCSname", "DPLuid", "DPLname", 
                  "DPLtype", "CMAPuid", "CMAuid", "CMAname", "CMAtype", "CTuid", 
                  "CTcode", "CTname", "POPCTRRAPuid", "POPCTRRAuid", 
                  "POPCTRRAname", "POPCTRRAtype", "POPCTRRAclass")
  )

  # Read raw 2011 attribute file 
  attribute_file_db11_2011_RAW <- 
    read_fwf("Data/attribute_file_db11_2011_RAW.txt", col_positions, skip = 1)
  
  # Rename variables and keep those we need
  attribute_file_db11_2011_REDUCED <- attribute_file_db11_2011_RAW |>
    select(da_id_11 = "DAuid",
           db_id = "DBuid",
           db_pop = "DBpop2011",
           pr_id = "PRuid",
           pr_name = "PRname",
           cd_id = "CDuid",
           csd_id = "CSDuid",
           csd_name = "CSDname",
           sactype = "SACtype",
           cma_id = "CMAuid",
           ct_id = "CTuid") |>
    
    # Save da_pop as a numeric variable
    mutate(db_pop = as.numeric(db_pop))
  
  # Collapse attribute file to DA level
  attribute_file_da11_2011_MASTER <- attribute_file_db11_2011_REDUCED |>
    group_by(da_id_11) |>
    summarise(da_pop_11 = sum(db_pop, na.rm = TRUE),
              pr_id_11 = first(pr_id),
              pr_name_11 = first(pr_name),
              cd_id_11 = first(cd_id),
              csd_id_11 = first(csd_id),
              csd_name_11 = first(csd_name),
              sactype_11 = first(sactype),
              cma_id_11 = first(cma_id),
              ct_id_11 = first(ct_id))
  
  # Count the number of unique DAs in 2011 attribute file
  n_distinct(attribute_file_da11_2011_MASTER$da_id_11)  # 56,204
  
  # Save the CSV file of the harmonized attribute file
  attribute_file_da11_2011_MASTER <- attribute_file_da11_2011_MASTER |>
    write_csv("Data/attribute_file_da11_2011_MASTER.csv")

  # 1.2. HARMONIZE 2016 ATTRIBUTE FILE AND CALCULATE POP COUNTS             ####
  # -------------------------------------------------------------------------- #
  
  # Add attribute file 2016 to R environment
  attribute_file_db16_2016_RAW <- 
    read_csv("Data/attribute_file_db16_2016_RAW.csv")
  
  # Rename variables
  attribute_file_db16_2016_REDUCED <- attribute_file_db16_2016_RAW |>
    select(da_id_16 = "DAuid/ADidu",
           db_id = "DBuid/IDidu",
           db_pop = "DBpop2016/IDpop2016",
           pr_id = "PRuid/PRidu",
           pr_name = "PRename/PRanom",
           cd_id = "CDuid/DRidu",
           csd_id = "CSDuid/SDRidu",
           csd_name = "CSDname/SDRnom",
           sactype = "SACtype/CSSgenre",
           cma_id = "CMAuid/RMRidu",
           ct_id = "CTuid/SRidu")
  
  # Collapse attribute file to DA level
  attribute_file_da16_2016_MASTER <- attribute_file_db16_2016_REDUCED |>
    group_by(da_id_16) |>
    summarise(da_pop_16 = sum(db_pop, na.rm = TRUE),
              pr_id_16 = first(pr_id),
              pr_name_16 = first(pr_name),
              cd_id_16 = first(cd_id),
              csd_id_16 = first(csd_id),
              csd_name_16 = first(csd_name),
              sactype_16 = first(sactype),
              cma_id_16 = first(cma_id),
              ct_id_16 = first(ct_id))
  
  # Count the number of unique DAs in 2016 attribute file
  n_distinct(attribute_file_da16_2016_MASTER$da_id_16)  # 56,590
  
  # Save the CSV file of the harmonized attribute file
  attribute_file_da16_2016_MASTER <- attribute_file_da16_2016_MASTER |>
    write_csv("Data/attribute_file_da16_2016_MASTER.csv")
  
  # 1.3. HARMONIZE 2021 ATTRIBUTE FILE AND CALCULATE POP COUNTS             ####
  # -------------------------------------------------------------------------- #
  
  # Add attribute file 2021 to R environment
  attribute_file_db21_2021_RAW <- 
    read_csv("Data/attribute_file_db21_2021_RAW.csv")
  
  # Rename variables and keep variables we need
  attribute_file_db21_2021_REDUCED <- attribute_file_db21_2021_RAW |>
    select(da_id_21 = "DAUID_ADIDU",
           db_id = "DBUID_IDIDU",
           db_pop = "DBPOP2021_IDPOP2021",
           pr_id = "PRUID_PRIDU",
           pr_name = "PRENAME_PRANOM",
           cd_id = "CDUID_DRIDU",
           csd_id = "CSDUID_SDRIDU",
           csd_name = "CSDNAME_SDRNOM",
           sactype = "SACTYPE_CSSGENRE",
           cma_id = "CMAUID_RMRIDU",
           ct_id = "CTUID_SRIDU")
  
  # Collapse attribute file to DA level
  attribute_file_da21_2021_MASTER <- attribute_file_db21_2021_REDUCED |>
    group_by(da_id_21) |>
    summarise(da_pop_21 = sum(db_pop, na.rm = TRUE),
              pr_id_21 = first(pr_id),
              pr_name_21 = first(pr_name),
              cd_id_21 = first(cd_id),
              csd_id_21 = first(csd_id),
              csd_name_21 = first(csd_name),
              sactype_21 = first(sactype),
              cma_id_21 = first(cma_id),
              ct_id_21 = first(ct_id))
  
  # Count the number of unique DAs in 2021 attribute file
  n_distinct(attribute_file_da21_2021_MASTER$da_id_21)  # 57,936
  
  # Save the CSV file of the harmonized attribute file
  attribute_file_da21_2021_MASTER <- attribute_file_da21_2021_MASTER |>
    write_csv("Data/attribute_file_da21_2021_MASTER.csv")

# 2. HARMONIZE CORRESPONDENCE FILES - 2016 AND 2021                         ####
# ---------------------------------------------------------------------------- #
# This step harmonizes the raw correspondence files of 2016 and 2021 to be able
# to join attribute files of two consecutive census for estimating intercensal 
# populations.
  
  # 2.1. HARMONIZE 2016 CORRESPONDENCE FILE                                 ####
  # -------------------------------------------------------------------------- #
  
  # Set directory of raw 2016 correspondence file
  raw_path <- "Data/correspondence_file_2016_RAW.csv"
  
  # Import correspondence file 2016
  correspondence_file_2016_RAW <- read_csv(raw_path)
  
  # Count unique DAs in the file - total obs = 57,300
  n_distinct(correspondence_file_2016_RAW$"DAUID2016/ADIDU2016")  # 56,590
  n_distinct(correspondence_file_2016_RAW$"DAUID2011/ADIDU2011")  # 56,204
  
  # ** The number of DA 2016 is the same as the number of DAs that exist 
  # in CASDOHI. **
  
  # Harmonize correspondence file
  correspondence_file_2016_HARM <- correspondence_file_2016_RAW |>
    
    # Select variables we need and rename them
    select(da_id_16 = "DAUID2016/ADIDU2016",
           da_id_11 = "DAUID2011/ADIDU2011",
           area_w = "DA_area_percentage/AD_pourcentage_superficie") |>
    
    # Recode area coverage to act as a weight
    mutate(area_w = area_w/100) |>
    
    # Save CSV version of the file
    write_csv("Data/correspondence_file_2016_HARM.csv")
  
  # 2.2. HARMONIZE CORRESPONDENCE FILE, 2021                                ####
  # -------------------------------------------------------------------------- #
  
  # Set directory of raw 2016 correspondence file
  raw_path <- "Data/correspondence_file_2021_RAW.csv"
  
  # Import correspondence file 2021
  correspondence_file_2021_RAW <- read_csv(raw_path)
  
  # Count unique DAs in the file - total obs = 59,551
  n_distinct(correspondence_file_2021_RAW$"DAUID2021_ADIDU2021")  # 57,936
  n_distinct(correspondence_file_2021_RAW$"DAUID2016_ADIDU2016")  # 56,590
  
  # The number of DA 2021 is the same as the number of DA that exist in CASDOHI.
  
  # Harmonize correspondence file
  correspondence_file_2021_HARM <- correspondence_file_2021_RAW |>
    
    # Select variables we need and rename them
    select(da_id_21 = "DAUID2021_ADIDU2021",
           da_id_16 = "DAUID2016_ADIDU2016",
           area_w = "DAAREAPRCNT_ADPRCNTSUP") |>
    
    # Recode area coverage to act as a weight
    mutate(area_w = area_w/100) |>
    
    # Save CSV version of the file
    write_csv("Data/correspondence_file_2021_HARM.csv")
  
# 3. JOIN HARMONIZED ATTRIBUTE AND CORRESPONDENCE FILES - 2016 AND 2021     ####  
# ---------------------------------------------------------------------------- #
  
  # 3.1 JOIN ATTRIBUTE AND CORRESPONDENCE - 2016                            ####
  # -------------------------------------------------------------------------- #  
   
  # Join attribute and correspondence files, 2016
  attribute_file_da11_2016_MASTER <- left_join(attribute_file_da16_2016_MASTER,
                                               correspondence_file_2016_HARM,
                                               by = "da_id_16")
  
  # Count unique DAs in the file - total obs = 57,300
  n_distinct(attribute_file_da11_2016_MASTER$"da_id_16")  # 56,590
  n_distinct(attribute_file_da11_2016_MASTER$"da_id_11")  # 56,204
  
  # Calculate population counts in year 2016 by 2011 DA
  pop_counts_da11_2016_MASTER <- attribute_file_da11_2016_MASTER |>
  
    # Calculate population in DA 11 by incorporating the percentage of the 
    # da 16 area they cover   
    mutate(da_pop_11 = da_pop_16 * area_w) |>
    
    # Collapse to da_11 to get population per da_11
    group_by(da_id_11) |>
    summarize(da_pop_16 = sum(da_pop_11)) |>
  
  # Save CSV
  write_csv("Data/pop_counts_da11_2016_MASTER.csv")
  
  # VALIDATION - Check and see if the 2016 population calculated for 2011 
  # geography match the one with 2016 geography to ensure accuracy.
  
  # Calculate 2016 Canada population by 2011 geography
  sum(pop_counts_da11_2016_MASTER$da_pop_16) # 35,151,728
  
  # Calculate 2016 Canada population by 2016 geography
  sum(attribute_file_da16_2016_MASTER$da_pop_16) # 35,151,728
  
  # 3.2 JOIN ATTRIBUTE AND CORRESPONDENCE - 2021                            ####
  # -------------------------------------------------------------------------- #  
  
  # Join attribute and correspondence files, 2021
  attribute_file_da16_2021_MASTER <- left_join(attribute_file_da21_2021_MASTER,
                                               correspondence_file_2021_HARM,
                                               by = "da_id_21")
  
  # Count unique DAs in the file - total obs = 59,551
  n_distinct(attribute_file_da16_2021_MASTER$"da_id_21")  # 57,936
  n_distinct(attribute_file_da16_2021_MASTER$"da_id_16")  # 56,590
  
  # Calculate population counts in year 2021 by 2016 DA
  pop_counts_da16_2021_MASTER <- attribute_file_da16_2021_MASTER |>
    
    # Calculate population in DA 16 by incorporating the percentage of the 
    # da 21 area they cover   
    mutate(da_pop_16 = da_pop_21 * area_w) |>
    
    # Collapse to da_16 to get population per da_16
    group_by(da_id_16) |>
    summarize(da_pop_21 = sum(da_pop_16)) |>

  # Save CSV
  write_csv("Data/pop_counts_da16_2021_MASTER.csv")

  # VALIDATION - Check and see if the 2021 population calculated for 2016 
  # geography match the one with 2021 geography to ensure accuracy.
  
  # Calculate 2021 Canada population by 2016 geography
  sum(pop_counts_da16_2021_MASTER$da_pop_21) # 36,991,981
  
  # Calculate 2021 Canada population by 2021 geography
  sum(attribute_file_da21_2021_MASTER$da_pop_21) # 36,991,981  
  
# 4. CALCULATE POPULATION COUNTS                                            ####
# ---------------------------------------------------------------------------- #                          
# In this step, we calculate the population for intercensal years based on 
# geographies of the former census year. The first step to do so is to link 
# the two datasets containing the population counts of two consecutive census.
# Next, we will calculate the intercensal population counts using linear
# interpolation method.
  
  # 4.1. CALCULATE POP COUNTS FOR 2011 TO 2015 BASED ON 2011 GEO            ####
  # -------------------------------------------------------------------------- #
  
  # Join 2011 and 2016 pop counts based on da_11
  pop_counts_da11_2011to2015_MASTER <- 
    left_join(attribute_file_da11_2011_MASTER,
              pop_counts_da11_2016_MASTER,
              by = "da_id_11") |>
    
    # Calculate and save difference in population between 2011 and 2016
    mutate(da_pop_dif = da_pop_16 - da_pop_11)
  
  # Calculate da-level population for years 2012 to 2015
  for (year in 12:15) {
    
    pop_counts_da11_2011to2015_MASTER <- pop_counts_da11_2011to2015_MASTER |>
      
    mutate(!!paste0("da_pop_", year) := da_pop_11 + (year - 11) * da_pop_dif/5)
  
    }
  
  # Save the standard geographies which we want to calculate pop counts for
  std_geo <- c("ct")
  
  # Save the years in which we want to calculate pop counts for
  years <- c("11", "12", "13", "14", "15")
  
  # Calculate population at larger geographies
  for (stg in std_geo) {
    for (year in years){
    
      pop_counts_da11_2011to2015_MASTER <- pop_counts_da11_2011to2015_MASTER |>
        group_by(across(all_of(paste0(stg, "_id_11")))) |>
        mutate(!!paste0(stg, "_pop_", year) := 
                 sum(.data[[paste0("da_pop_", year)]], na.rm = TRUE)) |>
        ungroup()
    
    }
  }
  
  # Save geographic levels that we estimated pop counts for
  geos <- c("da", "ct")
  
  # Round estimated intercensal pop counts to the nearest integer
  for (geo in geos) {
    for (year in years){
      
      pop_counts_da11_2011to2015_MASTER <- pop_counts_da11_2011to2015_MASTER |>
        mutate(!!paste0(geo, "_pop_", year) := 
                 round(.data[[paste0(geo, "_pop_", year)]]))   
    }
    
  }
    
  # Create a dataset for 2011 population counts
  pop_counts_da11_2011_RELEASE <- pop_counts_da11_2011to2015_MASTER |>
    select(ends_with("_11")) |>
    
    # Save csv to release
    write_csv("Data/pop_counts_da11_2011_RELEASE.csv")
  
  # Create a dataset for 2012 to 2015 population counts estimates
  pop_counts_da11_2012to2015_RELEASE <- pop_counts_da11_2011to2015_MASTER |>
    select( - ends_with("pop_11"), - da_pop_dif, - da_pop_16) |>
    
    # Save csv to release
    write_csv("Data/pop_counts_da11_2012to2015_RELEASE.csv")
  
  # 4.2. CALCULATE POP COUNTS BETWEEN 2016 AND 2021 BASED ON 2016 GEO       ####
  # -------------------------------------------------------------------------- #
  
  # Join 2016 and 2021 pop counts based on da_16
  pop_counts_da16_2016to2020_MASTER <- 
    left_join(attribute_file_da16_2016_MASTER,
              pop_counts_da16_2021_MASTER,
              by = "da_id_16") |>
    
    # Calculate and save difference in population between 2016 and 2021
    mutate(da_pop_dif = da_pop_21 - da_pop_16)
  
  # Calculate da-level population for years 2017 to 2020
  for (year in 17:20) {
    
    pop_counts_da16_2016to2020_MASTER <- pop_counts_da16_2016to2020_MASTER |>
      
      mutate(!!paste0("da_pop_", year) 
             := da_pop_16 + (year - 16) * da_pop_dif/5)
    
  }
  
  # Save the standard geographies which we want to calculate pop counts for
  std_geo <- c("ct")
  
  # Save the years in which we want to calculate pop counts for
  years <- c("16", "17", "18", "19", "20")
  
  # Calculate population at larger geographies
  for (stg in std_geo) {
    for (year in years){
      
      pop_counts_da16_2016to2020_MASTER <- pop_counts_da16_2016to2020_MASTER |>
        group_by(across(all_of(paste0(stg, "_id_16")))) |>
        mutate(!!paste0(stg, "_pop_", year) 
               := sum(.data[[paste0("da_pop_", year)]], na.rm = TRUE)) |>
        ungroup()
      
    }
  }
  
  # Save geographic levels that we estimated pop counts for
  geos <- c("da", "ct")
  
  # Round estimated intercensal pop counts to the nearest integer
  for (geo in geos) {
    for (year in years){
      
      pop_counts_da16_2016to2020_MASTER <- pop_counts_da16_2016to2020_MASTER |>
        mutate(!!paste0(geo, "_pop_", year) := 
                 round(.data[[paste0(geo, "_pop_", year)]]))   
    }
    
  }
  
  # Create a dataset for 2016 population counts
  pop_counts_da16_2016_RELEASE <- pop_counts_da16_2016to2020_MASTER |>
    select(ends_with("_16")) |>
    
    # Save csv to release
    write_csv("Data/pop_counts_da16_2016_RELEASE.csv")
  
  # Create a dataset for 2017 to 2020 population counts estimates
  pop_counts_da16_2017to2020_RELEASE <- pop_counts_da16_2016to2020_MASTER |>
    select( - ends_with("pop_16"), - da_pop_dif, - da_pop_21) |>
    
    # Save csv to release
    write_csv("Data/pop_counts_da16_2017to2020_RELEASE.csv")
  
  # 4.3. CALCULATE POP COUNTS FOR 2021 BASED ON 2021 GEO.                   ####
  # -------------------------------------------------------------------------- #
 
  # Create a dataset for 2021 population counts based on 2021 geographies
  pop_counts_da21_2021_RELEASE <- attribute_file_da21_2021_MASTER 
   
  # Save the standard geographies we need in a vector
  std_geo <- c("ct")
  
  # Calculate population at larger geographies using DA pop counts
  for (stg in std_geo) {
  
    pop_counts_da21_2021_RELEASE <- pop_counts_da21_2021_RELEASE |>
   
      group_by(across(all_of(paste0(stg, "_id_21")))) |>
      mutate(!!paste0(stg, "_pop_21") 
             := sum(da_pop_21, na.rm = TRUE)) |>
      ungroup()
    
  }
    
  # Save csv file to release
  write_csv(pop_counts_da21_2021_RELEASE, 
            "Data/pop_counts_da21_2021_RELEASE.csv")
  
  
  
  
  
  
  
  
  
  
  
  
  
  
  
  
  
  
  
  
  
  
  
  
  
```
